# Higher body mass index raises immature platelet count: evidence from Mendelian randomization analyses

**DOI:** 10.1101/2021.05.19.21257443

**Authors:** Lucy J. Goudswaard, Laura J. Corbin, Kate L. Burley, Andrew Mumford, Parsa Akbari, Nicole Soranzo, Adam S. Butterworth, Nicholas A. Watkins, Dimitri J. Pournaras, Jessica Harris, Nicholas J. Timpson, Ingeborg Hers

**Author notes:** **Corresponding author**: Ingeborg Hers.

## Abstract

A higher body mass index (BMI) is a recognised risk factor for thrombosis. Platelets are essential for haemostasis but also contribute to thrombosis when activated pathologically. We hypothesised that an increase in BMI may lead to changes in platelet characteristics, thereby contributing to increased thrombotic risk.

The effect of BMI on platelet traits measured by Sysmex XN-1000 was explored in 33388 UK blood donors from the INTERVAL study. Linear regression was used for observational analyses between BMI and platelet characteristics. Mendelian randomization (MR) was used to estimate a causal effect with BMI proxied by a genetic risk score. Follow-up analysis explored the relevance of platelet characteristics on whole blood platelet aggregation in a pre-operative cardiac cohort (COPTIC) using linear regression.

Observationally, higher BMI was positively associated with greater plateletcrit (PCT), platelet count (PLT), immature platelet count (IPC) and side fluorescence (SFL, a measure of mRNA content used to derive IPC). MR provided causal estimates for a positive effect of BMI on both SFL and IPC (IPC 0.06 SDs higher per SD higher BMI, 95% CI 0.006 to 0.12, P=0.03), but there was no strong evidence for a causal effect of BMI on PCT or PLT. The COPTIC study provided observational evidence for a positive association between IPC and whole blood platelet aggregation induced by adrenaline, TRAP-6 and ADP. Our results indicate that higher BMI raises the number of immature platelets, which is associated with greater whole blood platelet aggregation. Higher IPC could therefore contribute to obesity-related thrombosis.

**Essentials:**

- A higher body mass index (BMI) is associated with thrombotic disorders.
- We explored whether BMI is associated with platelets traits, key cells involved in thrombosis.
- We found causal evidence for higher BMI raising immature platelet count (IPC).
- Higher IPC is associated with enhanced platelet aggregation in a cardiac surgery cohort.

## Background

Obesity (body mass index ≥ 30 kg/m^2^) has nearly tripled worldwide in the last 45 years [1]. This is a major health concern as a higher body mass index (BMI) is an important risk factor for various noncommunicable disorders, including cardiovascular disease [2]. Clinical studies and genetic association studies have identified BMI as an independent risk factor for thrombotic disorders, including coronary heart disease and stroke [3–5].

Platelets are blood cells which are essential for the cessation of bleeding upon injury to a blood vessel and are involved in thrombosis and progression of cardiovascular disease [6]. When pathologically activated, platelets can aggregate to form thrombi thereby occluding arteries and triggering a myocardial infarction or stroke [6]. Platelet hyperactivity can be an indicator of those who may be at an increased risk of thrombosis [7]. There is evidence that people with a higher BMI have hyperactive platelets [8, 9], which could explain why a higher BMI is linked to an increase in thrombosis. Mechanisms of platelet hyperactivity are unclear, but it is possible that changes in platelet numbers, size and immaturity may play a role. Furthermore, an alteration in circulating metabolites or proteins induced by obesity could modulate the activation of platelets [10].

Platelet function can be directly assessed by various methods; these methods commonly include platelet aggregation experiments and/or using antibodies to detect platelet receptors that are expressed upon activation. One of the limitations of measuring platelet function directly is that these techniques are not widely available or readily standardized. Haematology analyzers are more commonly used to provide full blood counts, providing detailed readouts of platelet characteristics. Some of these characteristics have been reported to be indirect measures of platelet function [11]. For example, an increase in mean platelet volume (MPV) has been reported to be predictive of vascular mortality and ischaemic heart disease [12]. Another commonly measured platelet characteristic is the number of platelets in circulation (platelet count, PLT). An increase in platelet count has been reported to be associated with platelet hyperactivity [11] and there is evidence that higher platelet count is associated with ischaemic stroke [13].

Sysmex haematology analyzers are able to provide additional platelet characteristics, such as immature platelet fraction (IPF) and immature platelet count (IPC) [14]. Immature platelets (also known as reticulated platelets) are the youngest platelets in circulation and are detected based on having a greater forward scatter (FSC, an indicator of their size) and a greater side fluorescence (SFL, an indicator of their mRNA content) [15]. Higher levels of immature platelets are indicative of enhanced platelet production [16] and these platelets are believed to be more prothrombotic than older circulating platelets, with more dense granule release and increased P-selectin expression [15–17]. Higher immature platelet count is associated with adverse cardiovascular outcomes in patients with coronary artery disease [15] and reduced effectiveness of antiplatelet therapies [17, 18], suggesting that hyperactive immature platelets may contribute to adverse vascular events. Immature platelets have also been suggested to be less responsive to antiplatelet therapies such as prasugrel in acute coronary syndrome patients [17].

As these platelet characteristics provide potential information about platelet hyperactivity and given the increased thrombotic risk seen with adiposity, it is important to assess how adiposity affects platelet properties. Previous studies with modest samples sizes have implemented observational epidemiological methods to explore the effect of BMI on PLT, plateletcrit (PCT) and MPV. There is conflicting observational evidence regarding an association between BMI and PLT [19, 20], with some studies suggesting a positive association between BMI and MPV [21] and other studies reporting no such association [22]. Furthermore, as there is evidence that immature platelet production is increased in patients with metabolic syndrome and type II diabetes [23, 24], it is therefore important to explore whether BMI may be an independent predictor of this trait. It is currently unknown whether the influence of BMI on platelet properties and function are causal and independent of confounding effects.

In this study, we used data from the INTERVAL prospective cohort (N=33388) to explore the association between BMI and platelet traits. We combined observational and Mendelian randomization (MR) approaches to test the hypothesis that higher BMI leads to changes in platelet characteristics. Although observational studies can demonstrate associations between BMI and platelet characteristics, they cannot determine direct causality. To address the latter, we therefore employed MR, using a genetic risk score derived from single nucleotide polymorphisms (SNPs) associated with increased BMI. This allowed estimation of the causal effect of BMI on platelet traits, reducing the effect of confounding factors that are inherent to observational studies. To assess functional implications of BMI-platelet associations, a follow-up analysis was designed to explore the associations between platelet characteristics and whole blood aggregation in a cohort of cardiac surgery patients.

## Methods

### Study population

INTERVAL is a prospective cohort study that initially aimed to test the safety of reducing the time interval between donation of whole blood in 50000 participants [25]. Participants were over the age of 18 years, able to provide informed consent and free from a history of major disease. Participants were recruited between June 11^th^, 2012 and June 15^th^, 2014 from 25 National Health Service Blood and Transplant (NHSBT) centres across England. They filled out online questionnaires including self-reported height and weight, smoking status and alcohol consumption. Blood samples were also taken at baseline (before randomization within the study) where full blood counts were obtained. The study was approved by Cambridge East Research Ethics Committee. Permission for data access was provided by the Data Access Committee. Data contains sensitive content and requires permission to use therefore cannot be made publicly available. Access to data needs to be approved by the INTERVAL team www.intervalstudy.org.uk/more-information.

The present study was conducted on up to 33388 European ancestry participants living in the United Kingdom. These were the participants who were genotyped, had basic phenotype data and full blood count measures (**Figure 1**). Participants were mostly of European descent.

**Figure 1.**
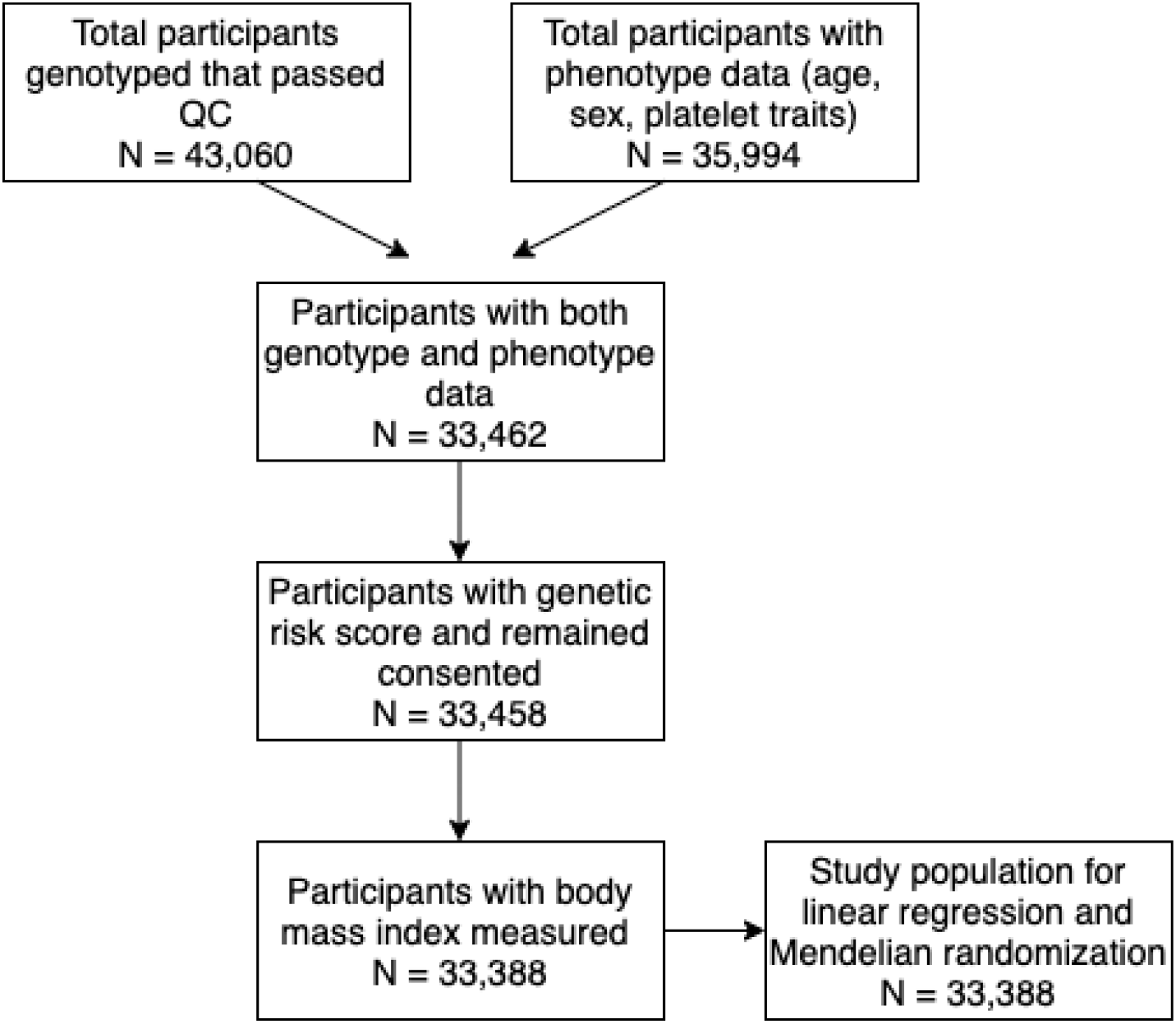
A STROBE diagram outlining participants included in the INTERVAL study (N=33388)

### Measurement of BMI and covariables

Participants self-reported their weight and height using online questionnaires. BMI was derived from their weight in kilograms by the square of their height in metres (kg/m^2^). BMI was rank normal transformed (*rntransform()* function from “moosefun” package https://github.com/hughesevoanth/moosefun/). Available covariables used in the analysis were age, sex, smoking status (in three categories of never, previous and current) and alcohol consumption (in four categories of rarely, less than once a week, 1-2 times a week and 3-5 times a week or most days). These covariables were chosen due to their plausible associations with both BMI and cardiovascular health [2], therefore it is important to adjust for these variables in the observational estimates.

### Measurement of platelet traits

Platelet parameters were measured using the Sysmex XN-1000 instrument [26]. This analyzer provides information on cell counts by using a combination of fluorescence (PLT-F) and impedance (I) flow cytometry. The PLT-F channel used a Fluorocell fluorescent dye (oxazine), whereas the impedance method uses electrical resistance to detect platelets. Platelet indices included in the current study, along with the raw units, are provided in **Table 1**. These measurements were pre-adjusted for technical covariates such as time between venipuncture and blood count, as well as instrument drift, seasonal and weekly variation [26]. Adjustment was performed with cyclic, thin-plate and P-splines in a generalized additive model (GAM) [27]. Resulting platelet trait values along with their units of measurement are reported in **Table 1**. These data were rank normal transformed to normalize the distribution of each trait. Therefore, each platelet index is measured in normalized standard deviation (SD) units.

**Table 1.** Platelet traits measured by Sysmex XN-1000.

| Short name | Full name | Derivation of platelet trait | Units | Mean (SD) |
| --- | --- | --- | --- | --- |
| P-LCR | Platelet large cell ratio | $\frac{\text{platelet number with volume} > 12 \text{ fL}}{\text{platelet number of ordinary volume}} \times 100$ | % | 36.9 (7.7) |
| H-IPF | High fluorescence immature platelet fraction | $\frac{\text{high fluorescence immature platelet number}}{\text{platelet count}} \times 100$ | % | 1.3 (1.0) |
| PLT (F) | Platelet count (PLT-F channel) | Total number of platelets per litre | 10<sup>9</sup>/L | 255 (57) |
| PLT (I) | Platelet count<br>(impedance<br>channel) | Total number of platelets per litre | 10<sup>9</sup>/L | 250 (55) |
| PCT | Plateletcrit | Volume percentage of blood occupied by platelets | % | 0.28 (0.06) |
| PDW | Platelet distribution<br>width | Standard deviation of platelet volume distribution,<br>estimated as width at 20% modal height of platelet<br>volume histogram | fL | 14.0 (1.8) |
| IPC | Immature platelet<br>count | Immature platelet count | 10<sup>9</sup>/L | 10.1 (5.0) |
| IPF | Immature platelet<br>fraction | $\frac{\text{immature platelet number}}{\text{platelet count}} \times 100$ | % | 4.2 (2.5) |
| MPV | Mean platelet<br>volume | Mean platelet volume | fL | 11.1 (1.0) |
| SFL | Side fluorescence | Side fluorescence measures the nucleic acid<br>(mRNA) content of the platelet | RFUs | 80.3 (5.0) |
| FSC | Forward scatter | Forward scatter is a measure of the size of the<br>platelet | RFUs | 53.0 (6.4) |
| SSC | Side scatter | Side scatter is a measure of the granularity of the<br>platelet | RFUs | 37.9 (2.9) |

### Genetic data and the association of the genetic risk score (GRS) with BMI

The genotyping of INTERVAL participants was performed using the Affymetrix GeneTitan® Multi-Channel (MC) Instrument. Quality control (QC) was performed as previously described [26]. Imputation was implemented using a combined 1000 Genomes phase-3-UK + 10K reference panel using the Practical Byzantine Fault Tolerance (PBFT) algorithm [26]. A genetic instrument for BMI was created by using 654 genetic variants available of the 656 variants which were independently associated with BMI (P<5×10^−8^) in a recent meta-analysis of GWAS of around 700,000 individuals of European descent [28]. Of these individuals, 450,000 were from UK Biobank and 250,000 were from the Genetic Investigation of Anthropometric Traits (GIANT) consortium. The weighted GRS was made using PLINK 2.0 [29]. The effect alleles and beta coefficients were extracted from the source GWAS. The weighted score was calculated by multiplying the dosage of each effect alleles by its effect estimate, summing these and dividing by the number of SNPs (654). The GRS therefore reflects the average per-SNP effect on BMI per individual.

### Statistical analysis

Analyses were performed using R version 3.4.2 [30]. To visualize correlation between outcome platelet variables, a correlation matrix was made using the pairwise Pearson correlation coefficients of the rank normal transformed data. A dendrogram was also created to show the hierarchical relationship between platelet traits (https://github.com/hughesevoanth/iPVs), where the height at which variables are joined is set at (1 – Pearson correlation coefficient (r)). To explore observational associations between BMI and platelet traits, linear regression models were used (*lm()* function from “stats” package). Two regression models were used: firstly, adjusting for age and sex and secondly, additionally adjusting for smoking and alcohol consumption as ordinal variables (**Error! Reference source not found**.). The results of the regression reflect the change in platelet traits (in normalized SD units) per normalized SD higher BMI (4.7 kg/m^2^). Only participants with all covariates were included in the linear regression models. The associations between potential covariables with both BMI and platelet traits were also explored using linear regression.

To understand properties of the GRS for BMI, the association between the GRS with both BMI and covariables were explored. The MR analysis was performed by using a two-stage least squares (2SLS) regression model (using *systemfit()* function from “systemfit” package [31], **Error! Reference source not found**.). The MR causal estimates reflect the change in platelet traits (in SD units) per SD increase in BMI. A Wu-Hausman test was performed to test for endogeneity between observational and MR estimates. Exact beta coefficients, confidence intervals and P values are provided throughout and guide strength of association.

### Follow-up analysis to explore the association between immature platelets and aggregation

Motivated by findings from the primary analyses described above, additional observational analyses were conducted using data from the COagulation and Platelet laboratory Testing in Cardiac Surgery (COPTIC) study. The COPTIC study was an observational, single centre cohort study of adults undergoing cardiac surgery at the Bristol Heart Institute with primary objective of examining the relationship between coagulation laboratory parameters and bleeding outcomes after surgery in 2541 participants [32]. This study was approved by the UK NHS Research Ethics Committee (09/H0104/53).

### COPTIC study variables

Age and sex were reported at baseline. Height and weight were obtained from medical notes. BMI was derived from weight and height (kg/m^2^). Smoking was reported as a categorical variable (0=never smoker, 1=ex-smoker for > 5 years, 2=ex-smoker for 1-5 years, 3=ex-smoker for 30 days-1 year, 4=current smoker). Platelet variables (PLT, MPV, IPF, IPC) were measured using the Sysmex XE-2100™ Automated Haematology System (Oxford, UK). Blood samples were taken pre-operatively into 3.2% sodium citrate vacutainers (BD Biosciences, Milton Keynes, UK). Platelet aggregation was measured using Multiplate multiple electrode aggregometry (MEA) (Roche, Rotkreuz, Switzerland), which detects change in electrical impedance when platelets aggregate on metal electrodes. Aggregation was determined by the area under the curve (AUC) in response to platelet agonists including adrenaline (100 mg/mL), thrombin receptor activator peptide 6 test (TRAP-test), ADP-test and ASPI-test. Use of a combination of agonists therefore reflects activation of platelets via different pathways: adrenaline acts at the α2 adrenergic receptor, TRAP-6 acts at the protease-activated receptor-1 (PAR-1), ADP acts at the P2Y12 receptor and arachidonic acid at the Thromboxane A2 (TxA2) receptor.

### COPTIC statistical analysis

The COPTIC dataset in the current analysis included 2518 participants (23 out of 2541 participants did not consent for future research). For the current study, only those who were not on antiplatelet therapies (prasugrel, clopidogrel or aspirin) were included (N=655). Extreme outliers that were ± 5 SDs from the mean were removed, and exposures and outcomes were rank normal transformed. Linear regression was used to explore the association between immature platelet count and aggregation, adjusting for age, sex and smoking status.

## Results

### INTERVAL participant characteristics

Of INTERVAL participants included in the current study (N=33388), 50.2 % were female. The mean age was 45.3 years (SD of 14.2 years, **Table 2**). The mean BMI was 26.4 kg/m^2^ (SD of 4.7 kg/m^2^). The majority of participants were never smokers (58.9 %), with 33.3% and 7.8 % reported as previous and current smokers, respectively. Nearly a third (32.7 %) of participants reported drinking alcohol at least three times a week.

**Table 2.**
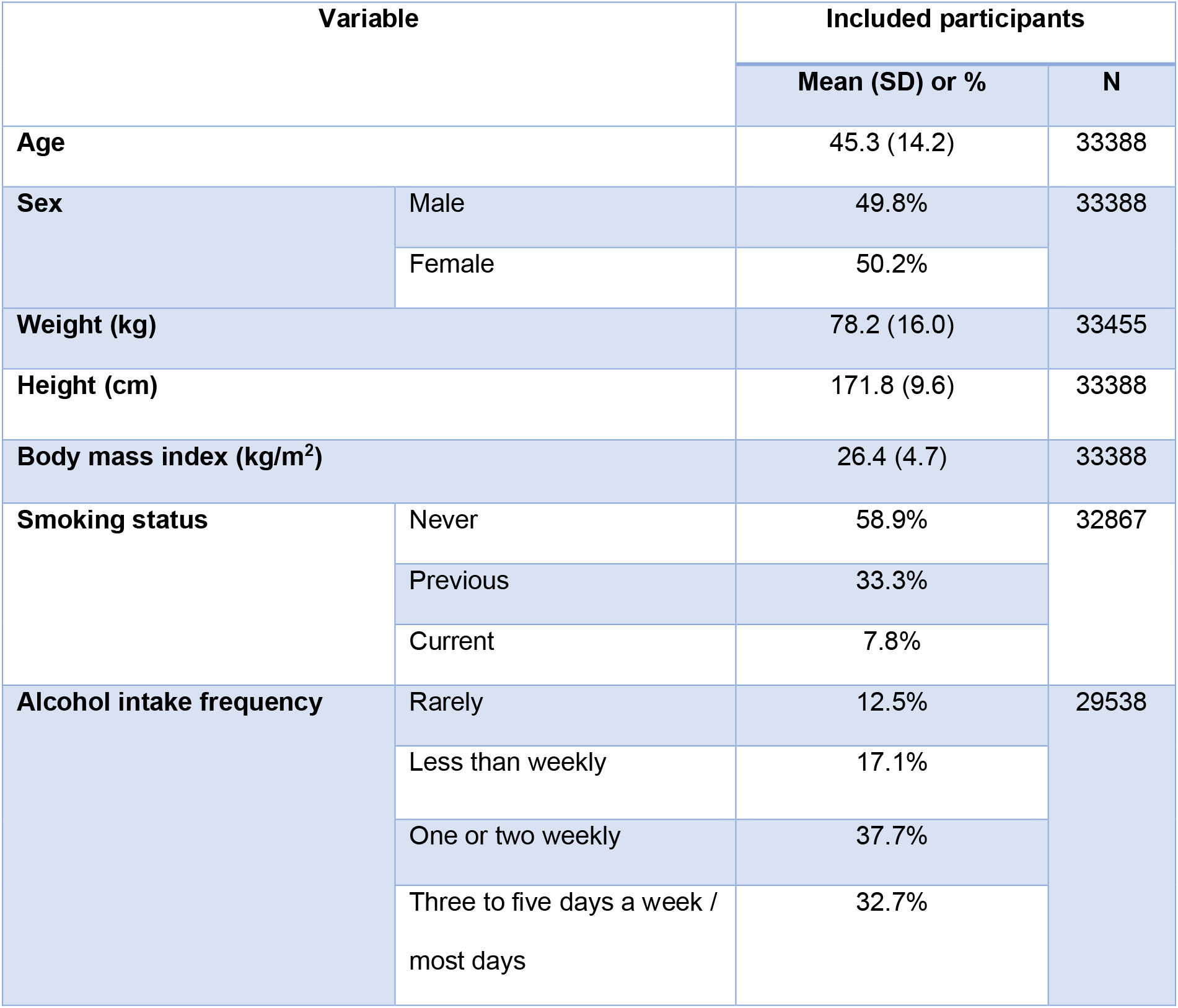
Characteristics of INTERVAL participants.

### Correlation between platelet traits in INTERVAL

The Sysmex XN-1000 haematology analyzer measures multiple platelet traits, however many of these traits are closely related measurements and therefore may not be completely independent. Indeed, platelet traits showed a high degree of correlation with each other (), in particular among similar measures. For example, measures of PLT (PLT I/F) and PCT, the latter a measurement of platelet mass, were highly positively correlated with each other but were weakly inversely correlated with other platelet measures. Measures of platelet maturity (IPF, IPC, H-IPF) were highly correlated with each other. In addition, platelet size variables (MPV, P-LCR and PDW) showed strong positive correlations with each other as well as with measures of immature platelets.

### Observational associations between BMI and platelet traits in INTERVAL

Linear regression was performed to determine whether a higher BMI was associated with a change in platelet characteristics. In the confounder adjusted linear regression (**Table 3**), BMI was positively associated with PCT (0.12 SD higher per SD higher BMI, 95% CI 0.11 to 0.13, P = 9.2 × 10^−88^) and platelet count (PLT (I) 0.11 SD higher per SD higher BMI, 95% CI 0.09 to 0.12, P = 1.0 × 10^−67^). The next strongest association with BMI was the association with SFL (0.06 SD higher per SD increase in BMI, 95% CI 0.05 to 0.07, P = 4.7×10^−23^). BMI also showed a positive association with IPC (0.06 SD higher per SD increase in BMI, 95% CI 0.05 to 0.08, P = 4.8×10^−22^). These results demonstrate that BMI is positively associated with PCT, PLT (I), SFL and IPC in this population. These estimates were very similar to the age and sex only adjusted estimates (**Supp Table 1**).

**Table 3.** **Observational associations between BMI and platelet measures (adjusted for age, sex, smoking status and alcohol consumption)**

| Platelet trait | N | Beta coefficient | SE | P value | R<sup>2</sup> | Eta<sup>2</sup> | 95% CI | Lower 95% CI | Upper 95% CI |
| --- | --- | --- | --- | --- | --- | --- | --- | --- | --- |
| PCT | 24562 | 0.12 | 0.006 | 9.17E-88 | 0.133 | 0.107 | 0.012 | 0.11 | 0.13 |
| PLT (I) | 26044 | 0.11 | 0.006 | 1.03E-67 | 0.096 | 0.081 | 0.012 | 0.09 | 0.12 |
| PLT (F) | 23921 | 0.10 | 0.006 | 5.62E-57 | 0.094 | 0.082 | 0.012 | 0.09 | 0.11 |
| SFL | 26082 | 0.06 | 0.006 | 4.73E-23 | 0.020 | 0.000 | 0.012 | 0.05 | 0.07 |
| IPC | 23909 | 0.06 | 0.007 | 4.84E-22 | 0.008 | 0.001 | 0.013 | 0.05 | 0.08 |
| PDW | 24563 | 0.03 | 0.007 | 5.21E-05 | 0.006 | 0.004 | 0.013 | 0.01 | 0.04 |
| FSC | 26082 | 0.02 | 0.006 | 0.01 | 0.026 | 0.001 | 0.012 | 0.01 | 0.03 |
| H-IPF | 26068 | -0.02 | 0.006 | 0.01 | 0.006 | 0.005 | 0.012 | -0.03 | -0.01 |
| IPF | 23909 | 0.01 | 0.007 | 0.04 | 0.008 | 0.007 | 0.013 | 0.00 | 0.03 |
| P-LCR | 27015 | 0.01 | 0.006 | 0.19 | 0.003 | 0.000 | 0.012 | 0.00 | 0.02 |
| **SSC** | 26082 | 0.01 | 0.006 | 0.28 | 0.040 | 0.000 | 0.012 | -0.01 | 0.02 |
| **MPV** | 24559 | 0.01 | 0.007 | 0.37 | 0.007 | 0.000 | 0.013 | -0.01 | 0.02 |

Beta coefficient is the change in platelet measure in SDs per normalized SD increase in BMI. PCT = plateletcrit, PLT (I) = platelet count (impedance channel), PLT (F) = platelet count (PLT-F channel), SFL = side fluorescence, IPC = immature platelet count, PDW = platelet distribution width, FSC = forward scatter, H-IPF = high fluorescence immature platelet fraction, IPF = immature platelet fraction, P-LCR = platelet large cell ration, SSC = side scatter, MPV = mean platelet volume. $\eta^2$ is the proportion of variance explained by the platelet trait in an ANOVA, whereas adjusted R squared is the variance explained by all the predictor variables in the regression model.

### Associations of covariables with BMI and platelet traits in INTERVAL

The association of covariables (age, sex, smoking and alcohol) with BMI were evaluated. Included covariables in the analysis showed associations with BMI (**Supp Table 2**). Males had a higher BMI than females (0.18 SD, 95% CI 0.16 to 0.21, P = 9.64×10^−64^). Age was positively associated with BMI (0.011 SD higher per year older, 95% CI 0.010 to 0.012, P = 1.11×10^−182^). Alcohol showed an inverse association with BMI and smoking showed a weak positive association with BMI.

Higher age was generally inversely associated with platelet measures (**Supp Table 3)**. Males had a lower platelet count and plateletcrit compared to females (PLT-F was 0.57 lower 95% CI −0.60 to −0.55, P = 9.9×10^−324^, **Supp Table 4**). Weak associations were detected between smoking and platelet traits such as a positive association between smoking status and immature platelets (**Supp Table 5**). Higher alcohol consumption also showed inverse associations with measures of plateletcrit and platelet count (**Supp Table 6)**.

**Table 4.** Association between genetic risk score for BMI with both exposure and covariables.

| Outcome | N | Beta coefficient | SE | P value | R<sup>2</sup> | F statistic |
| --- | --- | --- | --- | --- | --- | --- |
| **BMI (SDs)** | 33388 | 0.69 | 0.018 | 2.3E-312 | 0.042 | 1458.4 |
| **Age (years)** | 33388 | -1.46 | 0.26 | 2.2E-08 | 0.001 | 31.3 |
| **Sex (1 = female, 2 = male)** | 33388 | -0.01 | 0.009 | 4.7E-01 | -1.4E-05 | 0.5 |
| **Smoking (1= never, 2 = previous, 3 = current)** | 32867 | 0.05 | 0.012 | 9.2E-05 | 0.0004 | 15.3 |
| **Alcohol consumption (1 = Rarely , 2 = Less than weekly, 3 = One or two weekly, 4 = 3-5 weekly or every day)** | 29538 | -0.15 | 0.02 | 2.9E-15 | 0.002 | 62.4 |

*Beta coefficient is the change in outcome variable per unit increase in the genetic risk score for BMI*

**Table 5.**
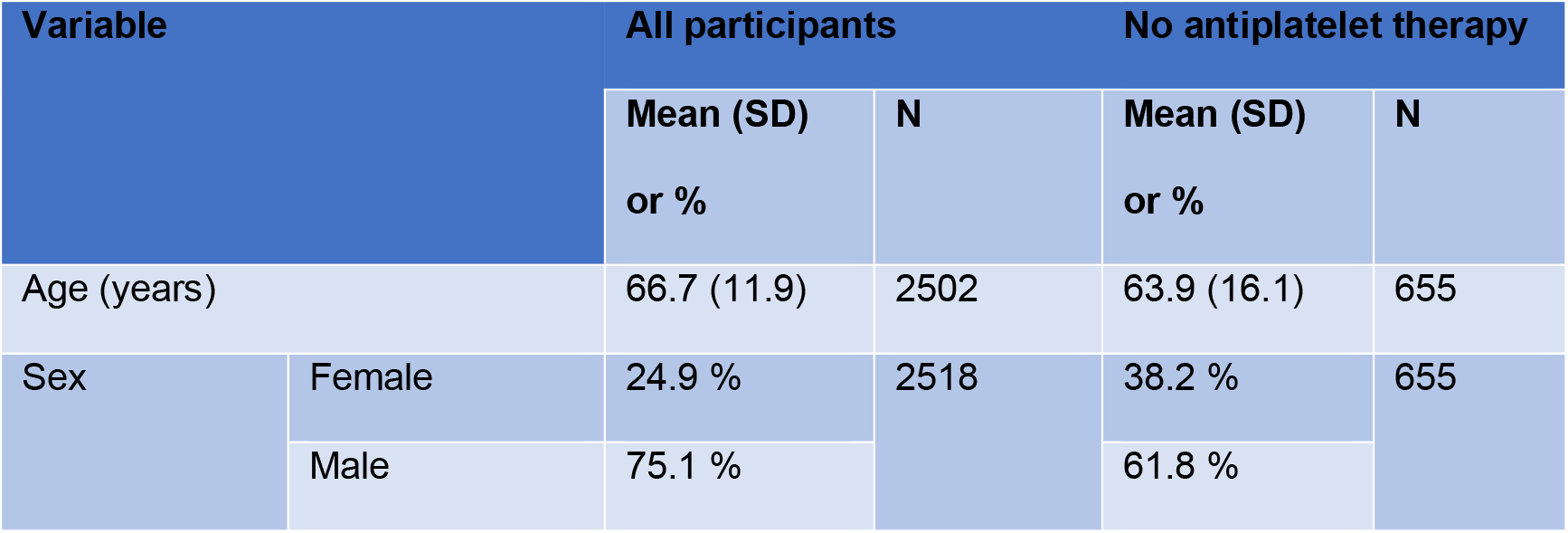
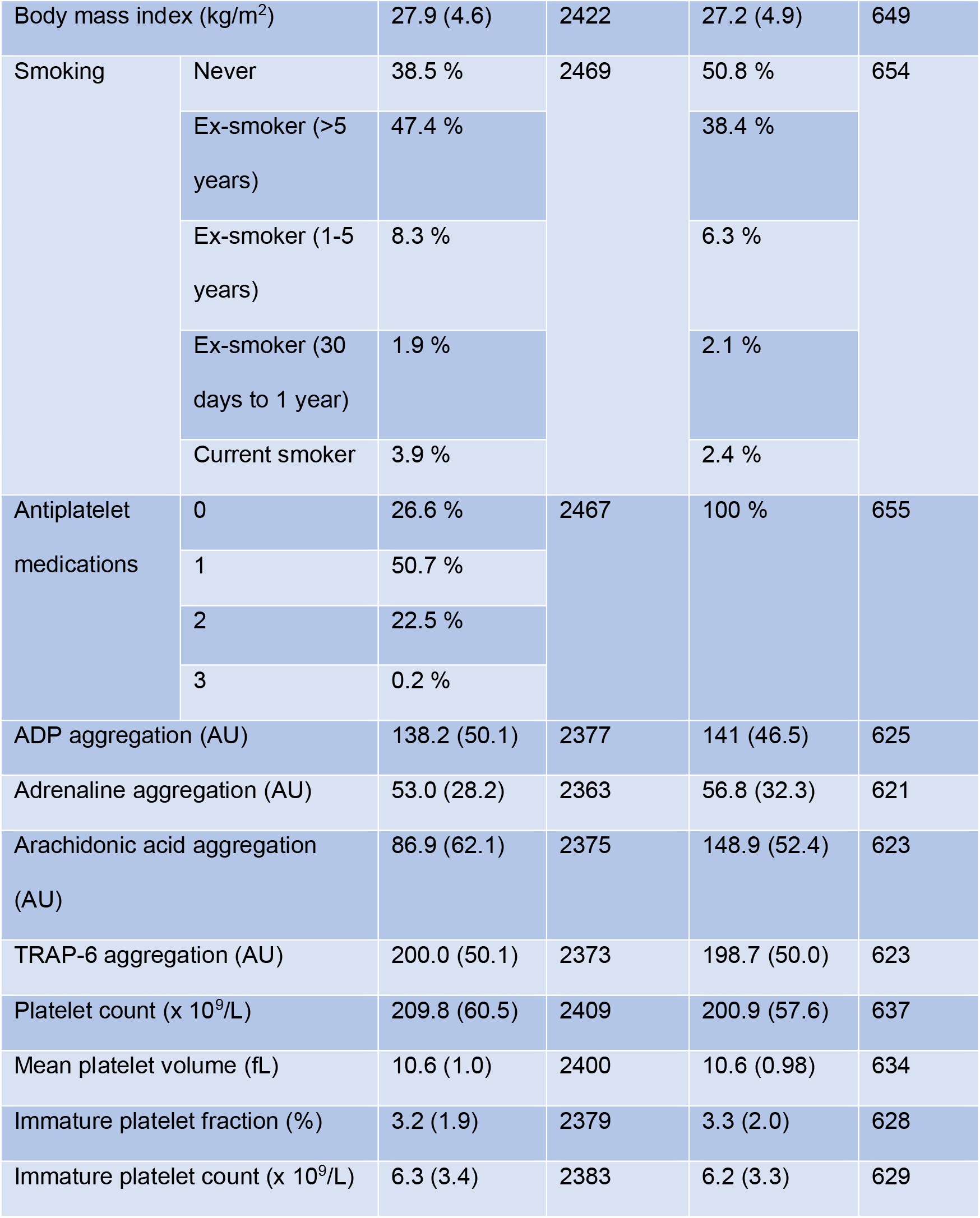
Characteristics of COPTIC study participants.

### GRS for BMI associations with BMI and covariables

To estimate the causal effect of BMI on platelet characteristics, MR was performed. The GRS for BMI (the average per SNP effect on BMI) showed a normal distribution (mean 0.08, SD 0.30, range −1.15 to 1.34) and was positively associated with BMI to the degree expected (R^2^ = 0.042, P = 2.3×10^−312^, F=1458, **Error! Reference source not found**.). The GRS did not associate with sex, however weak associations were detected with alcohol consumption, age, and smoking status. The amount of variance explained by the GRS for BMI on any covariable included did not exceed R^2^ = 0.002 (**Error! Reference source not found**.). As the GRS associates with BMI but does not strongly associate with the measured covariables, it is thus a valid instrument to perform MR.

### Mendelian randomization estimates for the association between BMI and platelet traits

In the MR analyses, BMI was associated with fewer traits than in the observational analysis (**Supp Table 7, Figure 4**). The causal estimate for the effect of BMI on SFL was 0.08 SDs per SD increase in BMI (95% CI 0.03 to 0.14, P = 0.003). This estimate was of larger magnitude than the observational estimate. The causal estimate for BMI and IPC was 0.06 SDs per SD increase in BMI (95% CI 0.006 to 0.12, P = 0.03), a similar magnitude of effect to the observational estimate. In the MR analysis, unlike in the observational analysis, the causal estimate did not suggest an effect of BMI on either PCT or PLT. MR estimates did not provide evidence for associations between BMI and other platelet variables. The point estimates were consistently positive, aligning with the positive observational associations seen across the platelet traits. Across all but one (H-IPF) of the platelet phenotypes we did not observe differences in directionality of effect comparing observational effects and causal effects predicted by Mendelian randomization (**Figure 4**).

**Figure 2.**
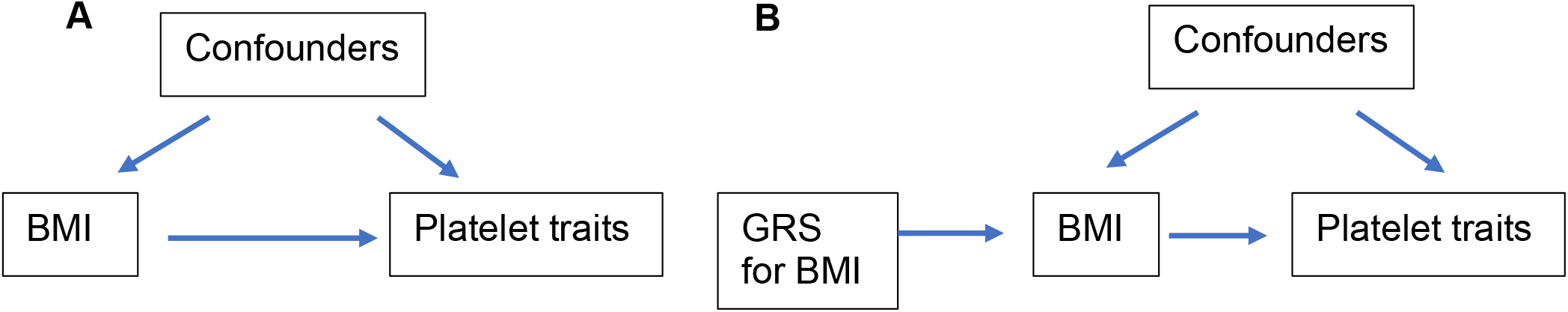
Schematic of linear regression and Mendelian randomization analyses. **A)** Linear regression assesses the association between BMI (exposure) and platelet traits (outcome), with adjustment for potential confounders. **B)** Under the assumptions of Mendelian randomization (MR), the genetic risk score (GRS) for body mass index (BMI) does not associate with confounding variables, and if there is a causal association, the GRS should only associate with platelet outcomes through its association with body mass index.

**Figure 3.**
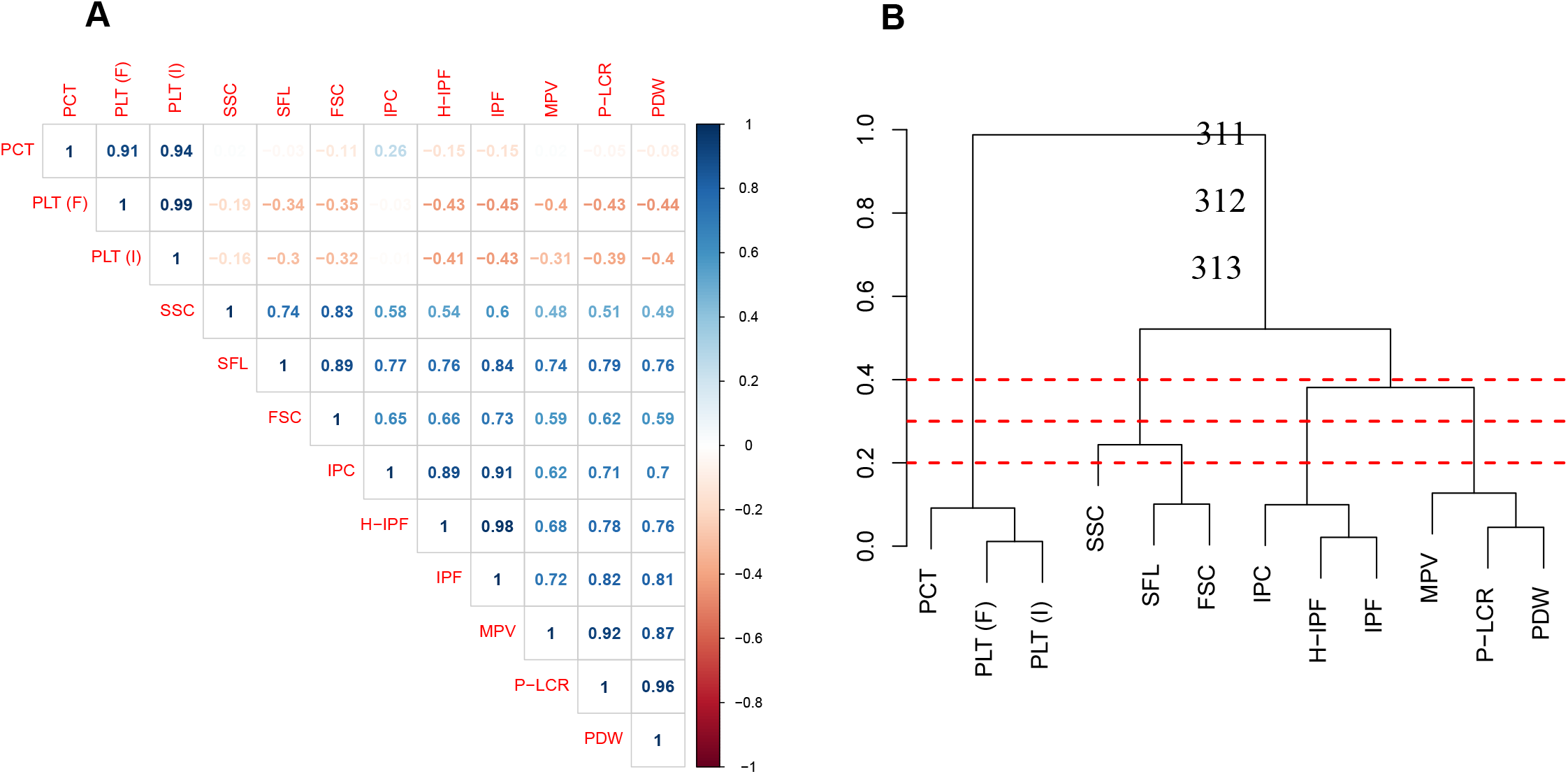
Correlation matrix and dendrogram of the relationship between platelet traits. **A)** Correlation coefficients are provided within the matrix. Dark blue indicates a correlation coefficient (r) of 1, with dark red indicating a correlation coefficient of −1. **B**) A dendrogram showing the hierarchical relationship between platelet traits, where the height at which two platelet traits join is (1 – Pearson correlation coefficient (r)). PCT = plateletcrit, PLT (F) = platelet count (PLT-F channel), PLT (I) = platelet count (impedance channel), SSC = side scatter, SFL = side fluorescence, FSC = side scatter, IPC = immature platelet count, H-IPF = high fluorescence immature platelet fraction, IPF = immature platelet fraction, MPV = mean platelet volume, P-LCR = platelet large cell ratio, PDW = platelet distribution width. Dashed red lines indicate a height of 0.2, 0.3 and 0.4.

**Figure 4.**
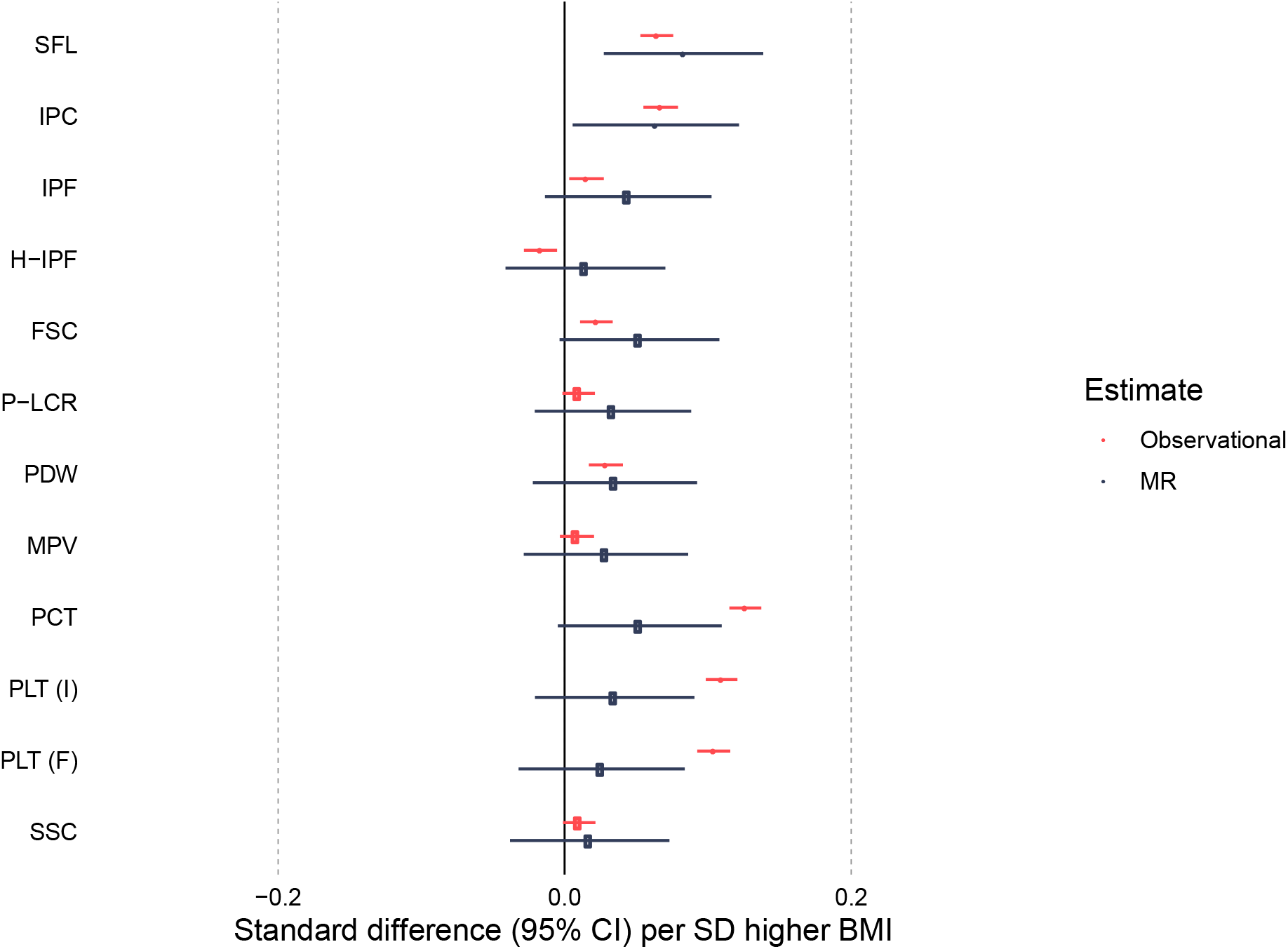
Forest plot of the unadjusted observational associations and MR estimates for BMI and platelet traits. Forest plot of the Mendelian randomization (MR) and unadjusted observational estimates for the effect of BMI on platelet traits. Estimate points are filled where P < 0.05. SFL = side fluorescence, IPC = immature platelet count, H-IPF = high fluorescence immature platelet function, FSC = forward scatter, P-LCR = platelet large cell ratio, PDW = platelet distribution width, MPV = mean platelet volume, PCT = plateletcrit, PLT (I) = platelet count (impedance channel), PLT (F) = platelet count (PLT-F channel), SSC = side scatter.

The Wu-Hausman test suggested that observational and MR estimates were similar for the majority of platelet traits (P > 0.05), except for measures of platelet count and plateletcrit (P < 0.001) (**Supp Table 7)**.

### Follow-up associations between IPC and whole blood aggregation in COPTIC

Given evidence for a causal effect of BMI on IPC in the MR analysis, we sought to evaluate the relationship between IPC and platelet activity as a biological parameter of clinical relevance. Whilst this analysis could not be conducted in INTERVAL due to a lack of suitable data, we were able to utilize data from the COPTIC study to address this question. The COPTIC study is a cohort of cardiac surgery patients, with samples taken pre-surgery. These participants have whole blood aggregation measured, therefore making it possible to determine associations between IPC and aggregation in a clinical setting.

### COPTIC participant characteristics

The total number of COPTIC participants was 2541. Of these, 2518 participants gave consent for future research (**Table 5**). Participants included in the analysis were those not on antiplatelet therapy (N=655). The majority of participants were male (61.8%), with a mean age of 63.9 years (SD of 16.1). Similar to the INTERVAL cohort, the mean BMI was in the overweight category (27.2 kg/m^2^ with a SD of 4.9 kg/m^2^). The majority of participants were either never smokers or ex-smokers for more than 5 years (89.2 %).

### Association between IPC and aggregation in COPTIC cohort

To determine the potential functional effects of variation in IPC, the COPTIC cohort was used to assess the observational association between IPC and whole blood platelet aggregation in response to a range of platelet agonists (**Figure 5A**). In participants who were not on antiplatelet therapy (up to N=655), there was evidence for a positive association between IPC and aggregation induced by adrenaline (0.13 SD increase per SD increase in IPC, 95% CI 0.04 to 0.21, p=3.7×10^−3^), TRAP-6 (0.11 SD increase per SD increase in IPC, 95% 0.03 to 0.18, p=7.8×10^−3^) and ADP (0.08 SD per SD increase in IPC, 95% 0.01 to 0.0.15, p=0.04). As there was a high correlation among platelet traits (**A, Figure 5B**), the effect of MPV, PLT and IPF were also assessed. PLT was associated with all four measures of aggregation, with a larger effect estimate than IPC (**Figure 5A**). The finding that IPC does not correlate with PLT however suggests that the effect of IPC on platelet aggregation is independent of the effect of PLT on platelet aggregation (**A, Figure 5B**). Fitting PLT alongside IPC as an additional predictor of aggregation had little effect on the IPC effect estimate providing further evidence of independent contributions from the two traits (**Figure 5C**). Adjustment for PLT in the regression model for the association between IPF and aggregation provided estimates of a similar magnitude to that of IPC and aggregation.

**Figure 5.**
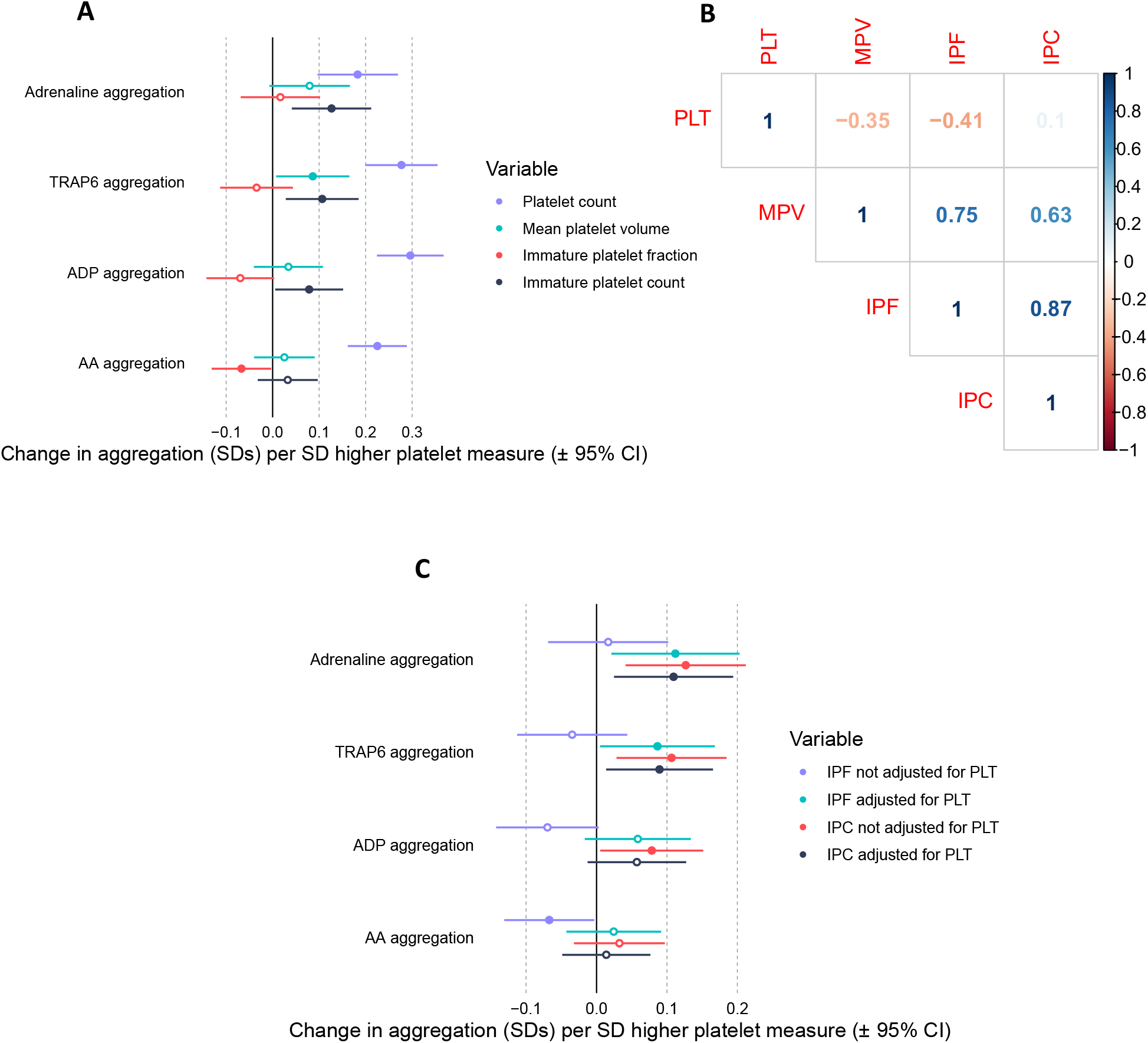
Association between platelet measures and aggregation in the COPTIC trial. A) Forest plot displaying the association between platelet measures (exposure) and aggregation (outcomes). B) Correlation matrix displaying the Pearson’s correlation coefficient (r) between platelet traits. C) Forest plot displaying the effect estimate for the association between immature platelet count (IPC) or immature platelet fraction (IPF) and measures of aggregation, with or without adjustment for platelet count (PLT). ADP = adenosine diphosphate, AA = arachidonic acid, MPV = mean platelet volume.

## Discussion

In this study we used data from 33388 healthy blood donors to study the effect of BMI on platelet phenotypes including measures of count, size and maturity. The combination of observational and MR estimates suggested a positive causal effect of BMI on SFL and IPC. The observational analysis revealed a strong association between BMI and both PLT and PCT, however the MR estimates did not provide evidence to support this association as causal. Observational analysis using data from a cardiac surgery cohort provided evidence for a positive association between IPC and aggregation induced by adrenaline, TRAP6 and ADP.

The observational and MR analyses in the current study provide evidence for a positive association between BMI and both IPC and SFL. As IPC is derived using SFL, the measure of mRNA content, these measures are highly correlated. Whilst few studies have explored the direct association between BMI and immature platelet measures, there is some evidence for increased immature platelets in subjects with metabolic syndrome (MS) and subjects with type II diabetes compared with control subjects [23, 24]. In these studies, participants with MS or type II diabetes also had a higher BMI, which could be a possible cause of this increase in immature platelets. Indeed, evidence from our analysis suggests that BMI, independent of metabolic syndrome or type II diabetes, may causally influence IPC and SFL.

As IPC is an indirect measure of platelet production [16], these results suggest that an increased BMI may lead to an increased production of platelets. It has been reported that immature platelets have increased thrombotic potential [16, 33] and may be predictive of cardiovascular events [15, 34], suggesting that an increase in the number of immature platelets may be of functional and clinical relevance. A follow-up analysis confirmed that higher IPC was associated with greater platelet aggregation independent of absolute platelet count within a cardiac surgery cohort. Positive associations were found between IPC and aggregation induced by adrenaline, TRAP-6 and ADP, suggesting general platelet hyperactivity with more immature platelets. A previous study in a coronary artery disease cohort also found a positive correlation between IPC and aggregation induced by arachidonic acid, collagen and ADP [33]. These findings suggest that IPC can be used as a proxy for platelet hyperactivity within a cohort with a history of cardiovascular disease. Measures of immature platelets are currently not considered in the clinic when prescribing antiplatelet therapy, however if a higher immature platelet count is demonstrative of newly produced, hyperactive platelets then this could be important in guiding dosage regimes to ensure sufficient platelet inhibition [17]. Observational studies have also found that patients with COVID-19 have elevated immature platelet counts and fraction, which could partly explain high rates COVID-19 induced thrombosis [35, 36]. Despite the association between BMI and IPC, there was less evidence for an effect of BMI on IPF. This lack of association may be because IPF is the proportion of immature platelets, therefore if someone had a higher number of immature platelets, but also a higher number of platelets overall (for example due to increased platelet lifespan), there would be no increase in the immature platelet fraction.

In general, previous studies have reported positive observational associations between BMI and both PLT and PCT [19]. More direct measures of body fat such as total fat mass, waist-hip ratio and waist circumference have also been reported to be positively associated with both PLT and PCT [20]; the observational evidence from the current study is in agreement with this. The MR analysis did not detect causal effects of BMI on measures of PLT/PCT, however the estimates were consistently positive. The small P values in the Wu-Hausman test for endogeneity for plateletcrit and platelet count variables support that the observational and MR estimates are different. Estimates could be biased due to confounding factors, reverse causation or other sources of bias. Possible routes of confounding could include stress, inflammation and nutrition, which could exert independent effects on both BMI and platelet count.

Both observational and MR estimates suggested that BMI is not associated with measures of platelet size, such as MPV and P-LCR. There are conflicting findings in the literature, with some studies suggesting that there is a positive association between BMI and MPV [21] and others showing no association [19, 20, 22]. The results of the current study suggest that associations seen previously between BMI and MPV are likely due to confounding of observational estimates. The lack of association between BMI and MPV may be surprising given a correlation coefficient of 0.62 between IPC and MPV. As BMI was positively associated with immature platelet count, and immature platelets tend to be larger, it may be surprising that BMI was not associated with measures of platelet size. However, immature platelets make up a small percentage of overall platelets (∼3-6 %) [15, 17], therefore the increase in immature platelets may not affect the overall median size of the whole platelet population.

Although this study suggests potential effects of BMI on platelet traits such as SFL and IPC, it does not provide mechanistic insight into how BMI exerts these effects. Previous studies have suggested that inflammation driven by adiposity can stimulate megakaryocyte proliferation, thereby increasing platelet numbers [20]. There is evidence that inflammatory mediators such as interleukin-6 (IL-6) could be one such factor [37]. Further study would be warranted to explore these mechanisms, as well as replicate the current findings, such as through independent population or clinical studies.

There are a few limitations to the study that should be recognised. Firstly, BMI was derived from self-reported height and weight. Although there is potential for this to bias observations, previous studies have found that self-reported BMI and BMI measured in the clinic are strongly associated [38]. The GRS also associates with BMI to the extent expected. Secondly, there may be other confounders which were not recorded within INTERVAL and therefore could not be accounted for in our models, such as, socio-economic position, which may affect both BMI and risk of thrombosis. Therefore, residual confounding of observational estimates cannot be ruled out. With respect to the observational analysis conducted in COPTIC, the sample size is modest, which may limit power to detect associations. Furthermore, this cohort required cardiac surgery and therefore it is possible that associations found may not be generalizable to the wider population. However, these findings do indicate that immature platelets may be a biomarker of platelet hyperactivity in patients with a history of cardiovascular disease.

Altogether, we show observational and MR evidence that an increased BMI is associated with an increase in number of immature platelets. Observational evidence indicates that higher immature platelet count is associated with enhanced aggregation in a cardiac surgery cohort. Together, these results indicate that higher BMI may enhance platelet function and thrombosis by increasing platelet production and immature platelet count.

## Data Availability

Data contains sensitive content and requires permission to use therefore cannot be made publicly available. Access to data needs to be approved by the INTERVAL team www.intervalstudy.org.uk/more-information.

## Addendum

LJG carried out the analysis and wrote the manuscript. LJC, KB and PA helped with statistical analysis and revising intellectual content. AM, NS, ASB, DJP helped with revising the intellectual content. NAW was involved in study design. JH was involved in the conduct, data analysis and linkage of the COPTIC study. NJT and IH were involved in concept and design of the study and revising the intellectual content.

## Acknowledgements

Participants in the INTERVAL randomised controlled trial were recruited with the active collaboration of NHS Blood and Transplant England (www.nhsbt.nhs.uk), which has supported field work and other elements of the trial. DNA extraction and genotyping was co-funded by the National Institute for Health Research (NIHR), the NIHR BioResource (http://bioresource.nihr.ac.uk) and the NIHR [Cambridge Biomedical Research Centre at the Cambridge University Hospitals NHS Foundation Trust]*. The academic coordinating centre for INTERVAL was supported by core funding from: NIHR Blood and Transplant Research Unit in Donor Health and Genomics (NIHR BTRU-2014-10024), UK Medical Research Council (MR/L003120/1), British Heart Foundation (SP/09/002; RG/13/13/30194; RG/18/13/33946) and the NIHR [Cambridge Biomedical Research Centre at the Cambridge University Hospitals NHS Foundation Trust]. A complete list of the investigators and contributors to the INTERVAL trial is provided in this reference [25]. The academic coordinating centre would like to thank blood donor centre staff and blood donors for participating in the INTERVAL trial. This work was supported by Health Data Research UK, which is funded by the UK Medical Research Council, Engineering and Physical Sciences Research Council, Economic and Social Research Council, Department of Health and Social Care (England), Chief Scientist Office of the Scottish Government Health and Social Care Directorates, Health and Social Care Research and Development Division (Welsh Government), Public Health Agency (Northern Ireland), British Heart Foundation and Wellcome. This work was also supported by the Wellcome Trust grant number 206194. *The views expressed are those of the authors and not necessarily those of the NHS, the NIHR or the Department of Health and Social Care.

William Astle provided guidance on platelet trait data measured by Sysmex. The COPTIC study was conducted within NIHR Programme Grant for Applied Research (RP-PG-0407-10384). Gavin J Murphy was the lead applicant and chief investigator of the COPTIC study. Zoe Plummer and Veerle Verheyden coordinated the COPTIC study and Kurtis Lee performed laboratory analyses.

## References

1. Blüher, M., Obesity: global epidemiology and pathogenesis. Nat Rev Endocrinol, 2019. 15(5): p. 288–298.

2. Bhaskaran, K., et al., Association of BMI with overall and cause-specific mortality: a population-based cohort study of 3·6 million adults in the UK. Lancet Diabetes Endocrinol, 2018. 6(12): p. 944–953.

3. Nordestgaard, B.G., et al., The effect of elevated body mass index on ischemic heart disease risk: causal estimates from a Mendelian randomisation approach. PLoS Med, 2012. 9(5): p. e1001212.

4. Dale, C.E., et al., Causal Associations of Adiposity and Body Fat Distribution With Coronary Heart Disease, Stroke Subtypes, and Type 2 Diabetes Mellitus: A Mendelian Randomization Analysis. Circulation, 2017. 135(24): p. 2373–2388.

5. Wolk, R., et al., Body mass index: a risk factor for unstable angina and myocardial infarction in patients with angiographically confirmed coronary artery disease. Circulation, 2003. 108(18): p. 2206–11.

6. Koupenova, M., et al., Thrombosis and platelets: an update. Eur Heart J, 2017. 38(11): p. 785–791.

7. Puurunen, M.K., et al., ADP Platelet Hyperreactivity Predicts Cardiovascular Disease in the FHS (Framingham Heart Study). J Am Heart Assoc, 2018. 7(5).

8. Barrachina, M.N., et al., GPVI surface expression and signalling pathway activation are increased in platelets from obese patients: Elucidating potential anti-atherothrombotic targets in obesity. Atherosclerosis, 2019. 281: p. 62–70.

9. Nardin, M., et al., Body Mass Index and Platelet Reactivity During Dual Antiplatelet Therapy With Clopidogrel or Ticagrelor. J Cardiovasc Pharmacol, 2015. 66(4): p. 364–70.

10. Goudswaard, L.J., et al., Effects of adiposity on the human plasma proteome: Observational and Mendelian randomization estimates. medRxiv, 2020: p. 2020.06.01.20119081.

11. Würtz, M., et al., Platelet aggregation is dependent on platelet count in patients with coronary artery disease. Thromb Res, 2012. 129(1): p. 56–61.

12. Slavka, G., et al., Mean platelet volume may represent a predictive parameter for overall vascular mortality and ischemic heart disease. Arterioscler Thromb Vasc Biol, 2011. 31(5): p. 1215–8.

13. Gill, D., et al., Genetically Determined Platelet Count and Risk of Cardiovascular Disease. Arterioscler Thromb Vasc Biol, 2018. 38(12): p. 2862–2869.

14. Unay Demirel, O., S. Ignak, and M.C. Buyukuysal, Immature Platelet Count Levels as a Novel Quality Marker in Plateletpheresis. Indian J Hematol Blood Transfus, 2018. 34(4): p. 684–690.

15. Ibrahim, H., et al., Association of immature platelets with adverse cardiovascular outcomes. J Am Coll Cardiol, 2014. 64(20): p. 2122–9.

16. Lev, E.I., Immature Platelets: Clinical Relevance and Research Perspectives. Circulation, 2016. 134(14): p. 987–988.

17. Bernlochner, I., et al., Impact of immature platelets on platelet response to ticagrelor and prasugrel in patients with acute coronary syndrome. Eur Heart J, 2015. 36(45): p. 3202–10.

18. Ibrahim, H., et al., Immature platelet fraction (IPF) determined with an automated method predicts clopidogrel hyporesponsiveness. J Thromb Thrombolysis, 2012. 33(2): p. 137–42.

19. Furuncuoğlu, Y., et al., How obesity affects the neutrophil/lymphocyte and platelet/lymphocyte ratio, systemic immune-inflammatory index and platelet indices: a retrospective study. Eur Rev Med Pharmacol Sci, 2016. 20(7): p. 1300–6.

20. Han, S., et al., Associations of Platelet Indices with Body Fat Mass and Fat Distribution. Obesity (Silver Spring), 2018. 26(10): p. 1637–1643.

21. Coban, E., et al., The mean platelet volume in patients with obesity. Int J Clin Pract, 2005. 59(8): p. 981–2.

22. Heffron, S.P., et al., Severe obesity and bariatric surgery alter the platelet mRNA profile. Platelets, 2018: p. 1–8.

23. Vaduganathan, M., et al., Platelet reactivity and response to aspirin in subjects with the metabolic syndrome. Am Heart J, 2008. 156(5): p. 1002.e1–1002.e7.

24. Mijovic, R., et al., Reticulated platelets and antiplatelet therapy response in diabetic patients. J Thromb Thrombolysis, 2015. 40(2): p. 203–10.

25. Di Angelantonio, E., et al., Efficiency and safety of varying the frequency of whole blood donation (INTERVAL): a randomised trial of 45 000 donors. Lancet, 2017. 390(10110): p. 2360–2371.

26. Astle, W.J., et al., The Allelic Landscape of Human Blood Cell Trait Variation and Links to Common Complex Disease. Cell, 2016. 167(5): p. 1415–1429.e19.

27. Akbari, P., et al., Genetic Analyses of Blood Cell Structure for Biological and Pharmacological Inference. bioRxiv, 2020: p. 2020.01.30.927483.

28. Yengo, L., et al., Meta-analysis of genome-wide association studies for height and body mass index in ∼700000 individuals of European ancestry. Hum Mol Genet, 2018. 27(20): p. 3641–3649.

29. Purcell, S., et al., PLINK: a tool set for whole-genome association and population-based linkage analyses. Am J Hum Genet, 2007. 81(3): p. 559–75.

30. R Core Team, R: A language and environment for statistical computing. R Foundation for Statistical Computing, Vienna, Austria. URL https://www.R-project.org/. 2019.

31. Henningsen, A. and J.D. Hamann, systemfit: A Package for Estimating Systems of Simultaneous Equations in R. 2007, 2007. 23(4): p. 40.

32. Mumford, A.D., et al., Near-patient coagulation testing to predict bleeding after cardiac surgery: a cohort study. Res Pract Thromb Haemost, 2017. 1(2): p. 242–251.

33. Grove, E.L., et al., Effect of platelet turnover on whole blood platelet aggregation in patients with coronary artery disease. J Thromb Haemost, 2011. 9(1): p. 185–91.

34. Freynhofer, M.K., et al., Platelet turnover predicts outcome after coronary intervention. Thromb Haemost, 2017. 117(5): p. 923–933.

35. Cohen, A., et al., Immature platelets in patients hospitalized with Covid-19. J Thromb Thrombolysis, 2020.

36. Klok, F.A., et al., Incidence of thrombotic complications in critically ill ICU patients with COVID-19. Thromb Res, 2020. 191: p. 145–147.

37. Kaser, A., et al., Interleukin-6 stimulates thrombopoiesis through thrombopoietin: role in inflammatory thrombocytosis. Blood, 2001. 98(9): p. 2720–5.

38. Nikolaou, C.K., C.R. Hankey, and M.E.J. Lean, Accuracy of on-line self-reported weights and heights by young adults. Eur J Public Health, 2017. 27(5): p. 898–903.

